# Robust Parental Preferences in Mental Health Screening in Youth From a Multinational Online Survey

**DOI:** 10.1101/2023.02.09.23285610

**Authors:** Mirelle Kass, Lindsay Alexander, Kathleen Moskowitz, Najé James, Giovanni Abrahão Salum, Bennett Leventhal, Kathleen Merikangas, Michael Peter Milham

**Affiliations:** Center for the Developing Brain, Child Mind Institute, New York, New York, USA; Graduate Program in Psychiatry and Behavioral Sciences, Universidade Federal do Rio Grande do Sul, Porto Alegre, Brazil; Section on Negative Affect and Social Processes, Hospital de Clínicas de Porto Alegre, Universidade do Rio Grande do Sul, Porto Alegre, Brazil; National Institute of Developmental Psychiatry for Children and Adolescents (INCT-CNPq), São Paulo, Brazil; Department of Psychiatry and Legal Medicine, Universidade Federal do Rio Grande do Sul, Porto Alegre, Brazil; Lifespan Informatics and Neuroimaging Center, Philadelphia, Pennsylvania, USA; The University of Chicago, Chicago, Illinois, USA; Genetic Epidemiology Research Branch, Intramural Research Program, National Institute of Mental Health, Bethesda, Maryland, USA; Nathan S. Kline Institute for Psychiatric Research, Orangeburg, New York, USA

## Abstract

**Importance:** Screening youth for mental disorders may assist in prevention, promote early identification, and reduce related lifetime impairment and distress.

**Objective:** The goal was to survey parents about their comfort and preferences for pediatric mental health screening, as well as factors associated with these preferences.

**Design:** The online survey was available July 11-14, 2021 on Prolific Academic. Analyses were conducted from November 2021 to November 2022.

**Setting:** Online survey.

**Participants:** The survey was administered to English-speaking parents with at least one 5-21-year old child at home. The sample included 972 parents, aged 21 and older, from the United States (*n*=265), United Kingdom (*n*=282), Canada (*n*=171), and Other Countries (*n*=254).

**Exposure(s):** None.

**Main Outcome(s)/Measure(s):** Parental preferences regarding the screening content, implementation preferences, and screener reviewing preferences of pediatric mental health screening were assessed in a novel survey. Mixed effects logistic models were employed to evaluate factors that influence parental comfort levels.

**Results:** Parents, aged 21 to 65 (*M*=39.4; 62.3% female), supported annual mental health screening for their child and preferred reviewing the screening results with professional staff (e.g., physicians). Parents preferred parent-report over child self-reports, though they were generally comfortable with both options. Despite slight variations based on country of residence, screening topic, and child’s age, parents were generally comfortable discussing all 21 topics. The greatest comfort was with sleep problems; the least comfort was with firearms, gender identity, suicidality, and substance use/abuse.

**Conclusions/Relevance:** Our data indicated that parents support annual parent- and child self-report mental health screening in primary care settings, but comfort levels differ according to various factors, such as screening topic. Parents preferred screening to occur in the healthcare office and to discuss screening results with professional staff. In addition to parental need for expert guidance, the growing awareness of child mental health needs highlights the importance of addressing mental health concerns early via regular mental health screenings.

**KEY POINTS:** *Question:* What are parents’ attitudes towards pediatric mental health screening in primary care settings?

*Findings:* The vast majority of parents surveyed online (*N=972)* expressed comfort with the screening of children for mental health concerns in the primary care setting. Variations in comfort were noted in relation to age of child and topics included. Parents expressed a preference for parent report over child report, as well as for reviewing screening results with professional medical staff. These findings were robust to the country of residence (e.g., United States, Canada, United Kingdom).

*Meaning:* Our findings document parental preferences that should be incorporated to enhance the feasibility of mental health screening in primary care settings.

## INTRODUCTION

The growing prevalence and burden of mental health disorders in pediatric populations have made clear the need for improved detection of mental disorders.^1–3^ In particular, early identification of youth mental disorders via universal screening is an increasingly actionable solution, with the potential to minimize the severity and progression of illness, mediate long-term impairment, and increase access to care,^4–7^ specially for common problems such as depression and anxiety.^8,9^ A growing literature has drawn attention to primary healthcare settings^1,10,11^ as a natural point of integration, noting both the breadth of screenings already included in “well-child” visits and the reality that most mental health difficulties are first discussed with primary care providers (PCPs).^12,13^ Consistent with these notions, recent work has found that mental health referrals from PCPs are preferred and result in a higher follow-up rate when compared to referrals from other parties.^14^ Accordingly, expert bodies, healthcare systems and local governments are increasingly promoting and building infrastructures to deploy mental health screening in primary care settings.^8,9,15–21^ Though, it is also important to maximize their acceptability. To date, much of the work around preferences and acceptability of screening has focused on medical staff.^22–25^ Here, we focus on the attitudes of parents and caretakers, which require careful attention in order to optimize implementation.

Preliminary findings suggest that parents are generally supportive of pediatric mental health screening,^26–30^ though, preferences do exist. For example, recent studies have proposed that both parents and medical staff prefer mental health screening to occur during annual, routine visits.^24,31^ While some studies have found that parents prefer to review their child’s screening results with staff that has medical expertise,^32,33^ there is variability based on the content of the screening instrument.^30^

Although these studies often show relatively high acceptance rates (75-85%), certain topics had significantly lower acceptance rates (e.g., 50.4%) and percentages of acceptance appear to drastically differ according to the topic.^30^ Attitudes towards a growing number of mental health topics are being assessed in the literature (e.g., suicidality,^25,30,34,35^ substance use,^24,30^ firearms,^26,30,36^ depression,^22,25,30,37^ attention-deficit hyperactivity disorder,^22,37^ anxiety,^22,37^ and gender identity^29^), though typically in isolation of one another^24,25,29,34,38^ - precluding a comprehensive picture.^22,26,30,37^ Additionally, these studies often focused on the attitudes of patients and medical staff rather than parents and caregivers; they rarely studied the impact of the report option for the topic in question (e.g., parent-report or a child self-report questionnaire). This is unfortunate because understanding parents’ comfort levels with mental health topics is essential for the development of effective screening procedures. In addition, parental acceptance of screening is likely to differ, depending on whether they or their child is having the conversation.^32^ Further, some have identified the limits of solely relying on parent “complaints’’ as a proxy for different types of problematic behavior in a child.^39–41^ Prior studies suggest that an individual’s country of residence may influence perceptions of mental disorders, in part due to cultural differences related to stigma and knowledge about resources for mental health.^42,43^ There is limited knowledge regarding parents’ comfort levels and preferences towards screening methods and content across international samples.

The present study examined comfort levels and preferences of parents towards mental health screening in pediatric primary care settings. In July 2021, a novel survey that incorporated previous questionnaires and research, along with input from experts, was administered to parents and caregivers from 19 different English-speaking countries through Prolific Academic, an online crowdsourcing platform. This survey assessed general views of pediatric screening, methods for administering screeners (setting, frequency, staff), screening content (21 topics), and the report option (parent-report versus child self-report). Preferences were compared across countries to examine the factors that influence parental preferences, comfort levels, and acceptability.

## METHOD

### Participant Recruitment

Data were collected July 11-14, 2021, through Prolific Academic (https://www.prolific.ac/) (PA), an online, crowdsourced survey recruitment service. PA participants have been shown to be more diverse and provide higher quality data than similar data collection platforms.^44^ We requested samples from the United States, the United Kingdom, and Canada, as well as other European and/or English-Speaking Countries (e.g., Australia). These countries were grouped because they did not have sufficient parent samples on the platform. We received 300 responses from the United States (US), United Kingdom (UK), and Other Countries samples and 197 responses for the Canadian sample. PA participants were required to be fluent in English, be a parent to one or more children (ages 5-21), and report about their oldest child in our age range, still living at home. There were no additional inclusion/exclusion criteria.

All data on PA were collected anonymously after agreeing to the Terms of Service; no additional informed consent required. IRB oversight was provided by the Advarra Institutional Review Board.

### Study Design and Measures

A novel survey was designed to address the goal of better understanding parental preferences towards mental health screening. Questions were based on a literature review of similar surveys, as well as feedback from primary care providers and mental health experts. The survey included five parts, explained below.

#### Background/Demographics

Parents were asked questions about their own and their child’s age, race/ethnicity, and gender identity. Additional questions addressed the family’s socioeconomic status, health insurance status, family history of mental illness, and frequency of doctor visits.

#### Mental Health Willingness

Respondents were asked to rate their agreement with 15 statements about mental health and learning disorders on a 6-point Likert scale (1=Disagree, 6=Agree). The 15 statements assessed willingness for discussions (e.g., “I am willing/able to discuss mental health with my child” and “I am willing/able to talk about my child’s learning difficulties with my family”) and perceptions of mental health (e.g., “It should be equally easy to talk about both mental health and physical health”).

#### Screening Administration Method

Participants were asked 7 questions to assess their preferred mental health screening setting. One item queried the desired frequency of screening (“monthly”, “quarterly”, “annually”, “never”). Five items assessed preferred screening setting(e.g., “in healthcare office, at annual well-child visit only,” and “at home, telehealth visit”) on a 6-point Likert scale (1=Disagree, 6=Agree). One multi-select checkbox item assessed parents’ preference regarding which staff member (physician, nurse, other health care provider, office staff, social worker, psychologist, counselor, teacher, and other) they would like to discuss their child’s mental health issues.

#### Screening Benefits and Feedback

4 items assessed parents’ opinions regarding the possible benefits of mental health screening. The listed benefits were “early detection of problems,” “early intervention,” “learning more about my child,” and “other.” Participants rated their agreement with each benefit on a 6-point Likert scale (1=Disagree, 6=Agree), and then were offered a free response option to suggest additional benefits. 4 items assessed parents’ preferences towards who completes the screener and their preference for receiving results and feedback.

#### Parental Comfort with Screening Topics

Parents’ comfort levels with 21 topics were assessed as a parent-report option and as a child self-report option. Topics included depression, autism, suicidality, neurodevelopmental disorders, firearms, gender identity, and social media use. Comfort levels were rated on 6-point Likert scales, with 6 indicating high comfort.

### Statistical Analysis

Data were collected from 1136 participants; we excluded 164 participants, 22 of whom did not report their child’s age, and 142 of whom had children outside of the age range of our inclusion criteria. The final sample included 972 parents with children aged 5-21 (*M*=11.1, *SD*=4.3). Parents were grouped by their country of residence to ensure adequate power for across-country analyses. The final sample included 265 US parents, 282 UK parents, 171 parents from Canada, and 254 parents from Other Countries. About 1% of participants did not complete the entire survey; their completion rates ranged from 70-87%.

Statistical analyses were conducted using R (lme4^45^, stats^46^). Descriptive statistics were determined for survey sections prior to more advanced analyses being conducted. For all analyses, a two-sided statistical significance cut-off of <.05 was applied. Benjamini-Hochberg correction^47^ was applied as appropriate. Linear and mixed-effect multivariate regression models were conducted to explore whether certain variables, or interaction of variables, significantly predicted parental comfort levels as a random variable.

## RESULTS

### Sample Characteristics

The final sample consisted of 972 parents (White: 75.3%; Female: 62.3%) aged 21 or older (*M*=39.4, *SD*=6.9) from the US (*n*=265), UK (*n*=282), Canada (*n*=171), or Other Countries (*n*=254). Children were between the ages of 5 and 21 (US: *M*=11.3 *SD*=4.2; UK: *M*=10.7, *SD*=4.4; Canada: *M*=11.4, *SD*=4.3; Other Countries: *M*=11.1, *SD*=4.4) The US, UK, and Canada samples were predominantly female (65.7%, 80.5%, 49.7%, respectively); the Other Countries sample was predominantly male (52.0%). The Other Countries sample included parents from 16 different countries. See Table 1 for demographic data.

**Table 1.**
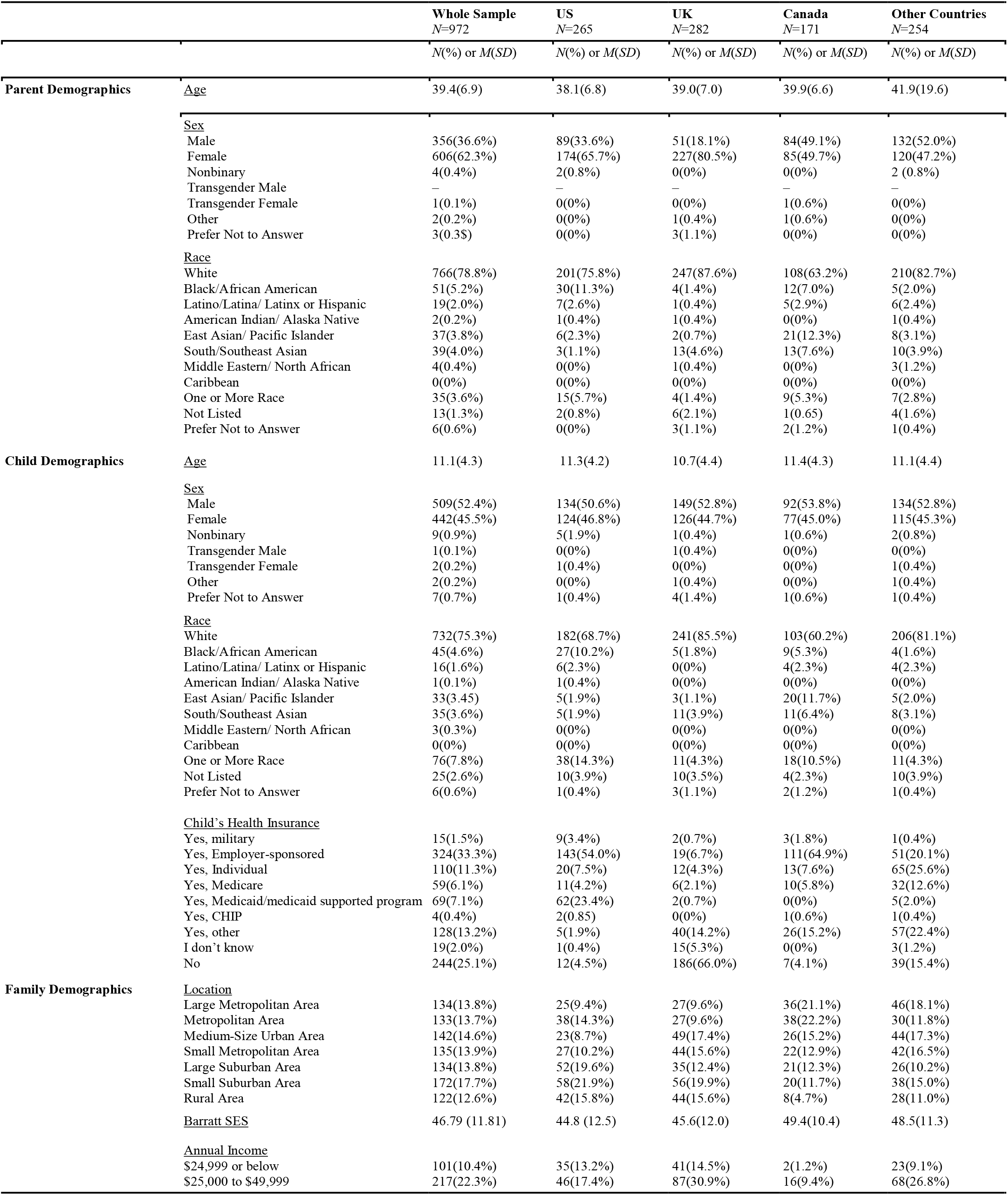
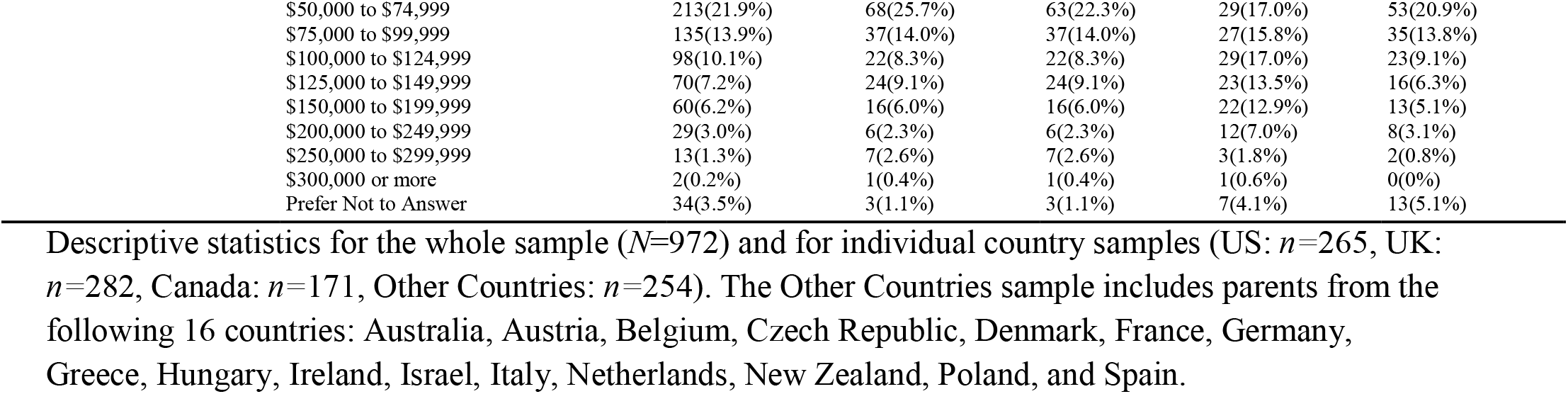
Sample Characteristics

### Parents Want Their Children Screened for Mental Health and Learning Problems

93% of parents (*n*=895) reported that they want their child screened for mental health problems at regular intervals. Annual screening was preferred by 65% of the sample (*n*=631), followed by quarterly (28%; *n*=226).

Across the entire sample, parents were most willing (yes/no) to speak with physicians (89.7%; *n*=872), followed by psychologists (76.4%; *n*=743). Only 5% (*n*=46) of parents were willing to discuss the screening results with general office staff (Figure 1). Figure 1 depicts the slight variances that were observed by country. When looking at specific data sources, the most notable variations across country samples were parents’ willingness to speak with teachers (US: 42.6%, UK: 61.7%, Canada: 46.2%, Other Countries: 44.5%) and social workers (US: 35.5%, UK: 37.2%, Canada: 49.7%, Other Countries: 26.4%). Over 65% of parents from each country sample expressed a willingness to talk about their child’s mental health with physicians and psychologists, whereas less than 8% of parents from each country were willing to review screener results with general office staff.

**Figure 1.**
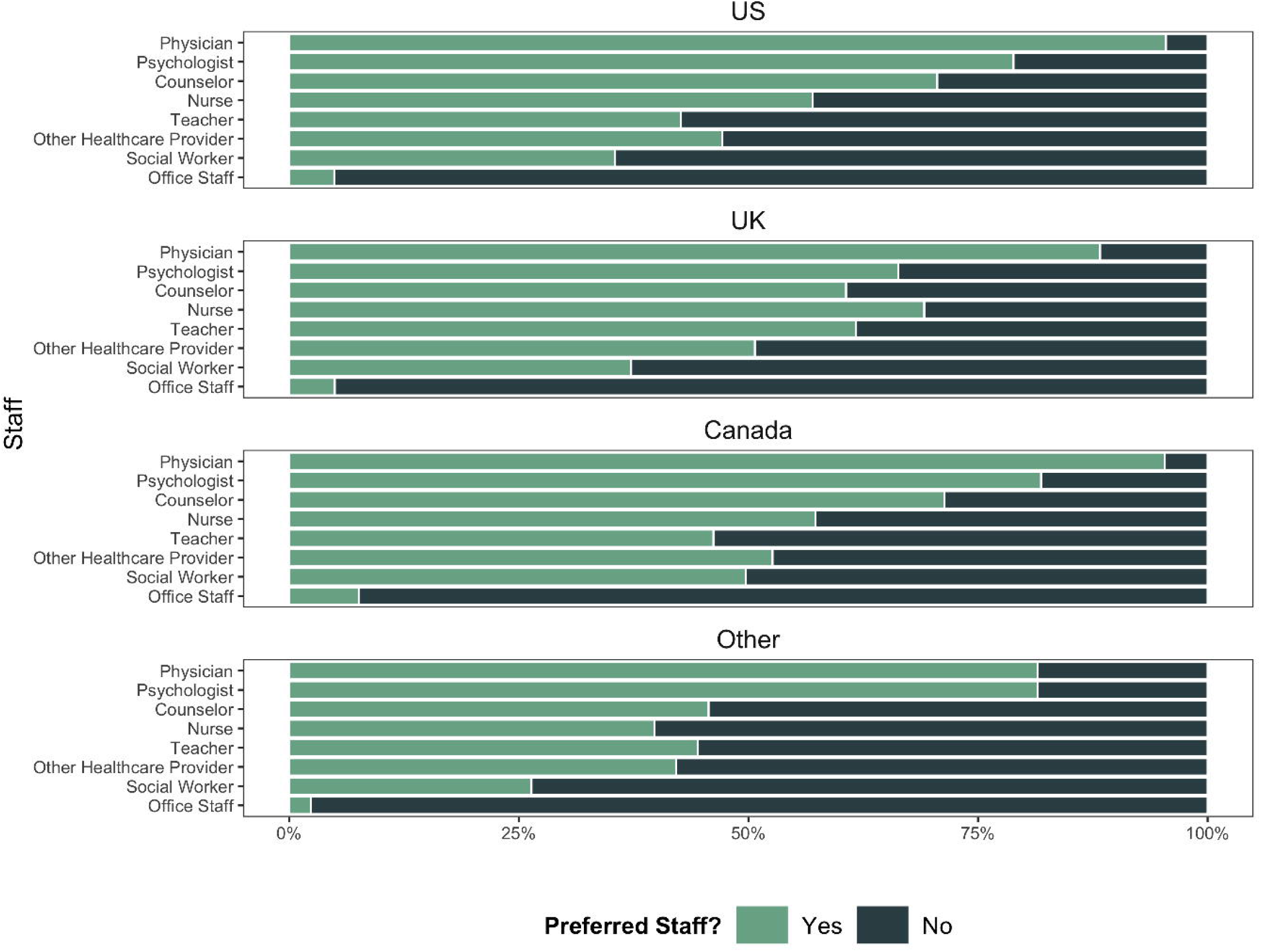
Parental Preferences for Reviewing Screening Findings for Each Country Sample: US, UK, Canada, and Other Countries. Parental preferences for reviewing their child’s screening results with 8 different staff members depicted in stacked bar plots by country sample. Parents were asked to report whether they were comfortable (“Yes”) or not comfortable (“No”) with each staff member.

Screening contexts and topics were assessed on a 6-point Likert scale, with 6 indicating the highest level of comfort. Figure 2 shows parents’ comfort level for five screening contexts. Parents indicated lower comfort with screening at home compared to in the healthcare office (*b*=-0.44, *p*<.001). Parents’ country of residence and the child’s age were accounted for in the regressions. Compared to the US sample, parents from Other Countries (*b*=-0.40, *p*<.001) and from the UK (*b*=-0.20, *p*<.001) reported decreased levels of comfort; no statistically significant difference was reported by parents from Canada. Higher comfort levels were associated with older children (*b*=0.02, *p*<.001).

**Figure 2.**
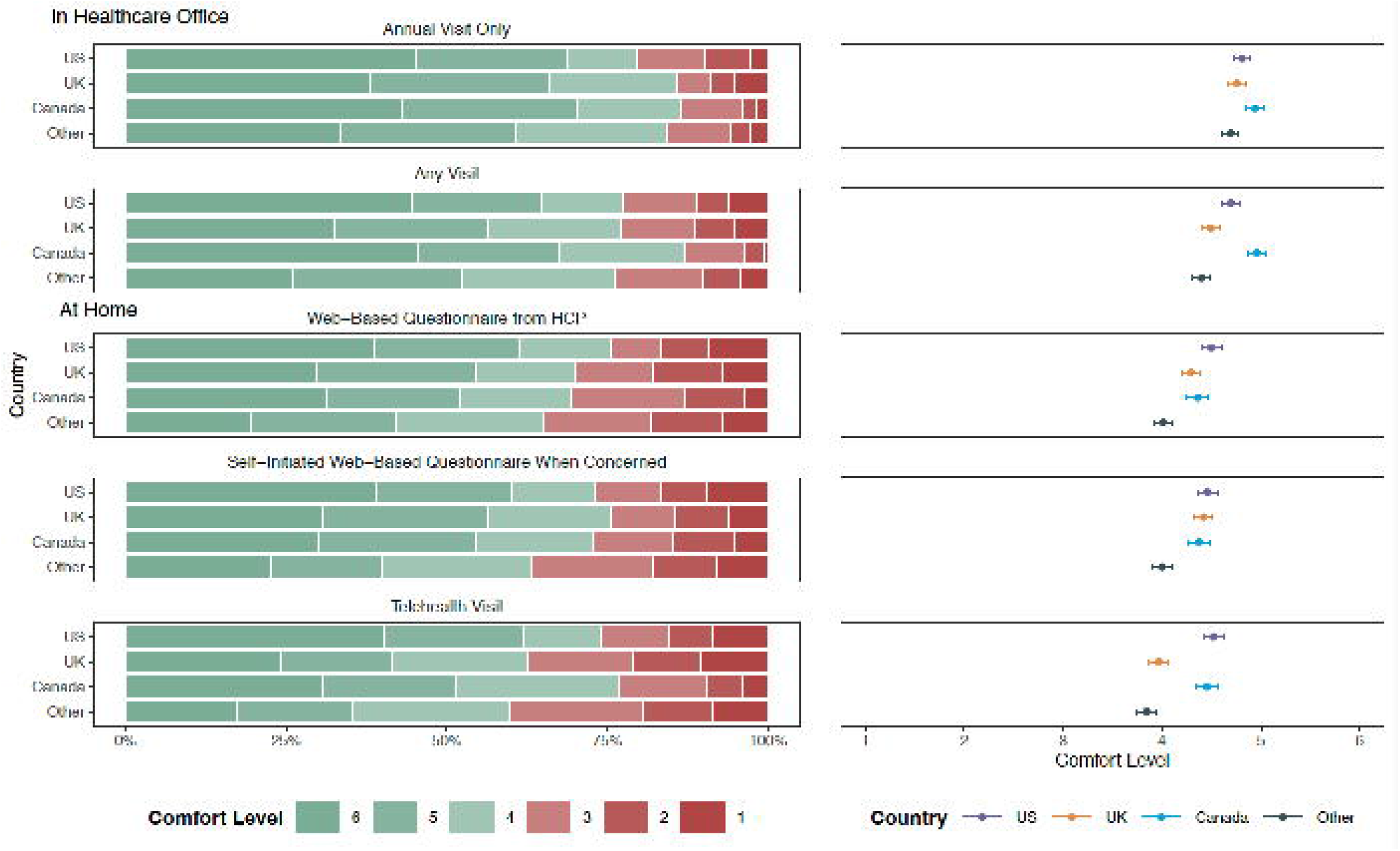
Parental Comfort Levels for Five Screening Administration Settings. Frequency of and average parental comfort levels by country sample for 5 screening settings are depicted in stacked bar plots and scatter plots, respectively. Comfort levels ranged from 1 to 6, with 6 indicating great comfort

A regression model compared the three at-home administration contexts: self-guided web-based screeners (*M*=4.31, *SE*=0.05), telehealth visits (*M*=4.17, *SE*=0.05), and provider-guided web-based screeners (*M*=4.29, *SE*=0.05); as well as two in-office administration contexts: annual visits (*M*=4.79, *SE*=0.04) and any visit (*M*=4.61, *SE*=0.05). Compared to the context with the highest average comfort level (in office, annual visit), the model found statistically significant lower comfort for the four other contexts: in office, any visit (*b*=-0.18, *p*<.01), at home, self-guided web-based screener (*b*=-0.47, *p*<.001), at home, provider-guided web-based screener (*b*=-0.50, *p*<.01), and at home, telehealth visit (*b*=-0.62, *p*<.01).

A mixed effect regression model that included parents’ country of residence, the child’s age, each screening topic, and the respondent option, found that, on average, parents reported significantly decreased comfort with child self-report compared to parent-report screeners (*b*= -0.278, *SE* = 0.009, *p*<.001). Furthermore, for every year that the child grew older, parental comfort levels increased (*b*= 0.035, *SE* = 0.008, *p*<.001). Mixed effect regression models by each of the topics found that parents were significantly more comfortable with parent-report compared to child self-report for all 21 topics (*p*<.01- *p*<.001). All parent-versus child self-report findings remained significant after corrections using the Benjamini-Hochberg method^47^ (*p*<.01-*p*<.001).

As depicted in Figure 3, parental comfort levels with 21 screening topics differed by the topic itself and by the report option (parent-report versus child self-report). Average parental comfort levels ranged from 4.62 (*SE*=0.05) to 5.30 (*SE*=0.03). Topics on which parents were most comfortable reporting included child sleep problems (*M*=5.30, *SE*=0.03), covid-19 concerns (*M*=5.23, *SE*=0.04), digital media use (*M*=5.22, *SE*=0.04), social media use (*M*=5.21, *SE*=0.04), and learning concerns (*M*=5.20, *SE*=0.04). Parents were the least comfortable reporting on their child’s experience with substance use/abuse (*M*=4.78, *SE*=0.05), firearms (*M*=4.71, *SE*=0.05), gender identity (*M*=4.68, *SE*=0.05), and suicidal ideation (*M*=4.62, *SE*=0.05). For child self-report, average parental comfort levels on a 6-point Likert scale ranged from 4.13 (*SE*=0.06) to 5.08 (*SE*=0.04). On average, parents were the most comfortable with their child reporting on their digital media use (*M*=5.08, *SE*=0.04), sleep problems (*M*=5.08, *SE*=0.04), social media use (*M*=5.06, *SE*=0.04), covid-19 concerns (*M*=5.06, *SE*=0.04), and bullying (*M*=5.04, *SE*=0.04). The child self-report topics that parents were the least comfortable with were gender identity (*M*=4.34, *SE*=0.06), substance use/abuse (*M*=4.34, *SE*=0.06), firearms (*M*=4.25, *SE*=0.06), and suicidal ideation (*M*=4.13, *SE*=0.06). Some variations were observed in parental comfort of specific topics across country samples (eFigure 2 in Supplement).

**Figure 3.**
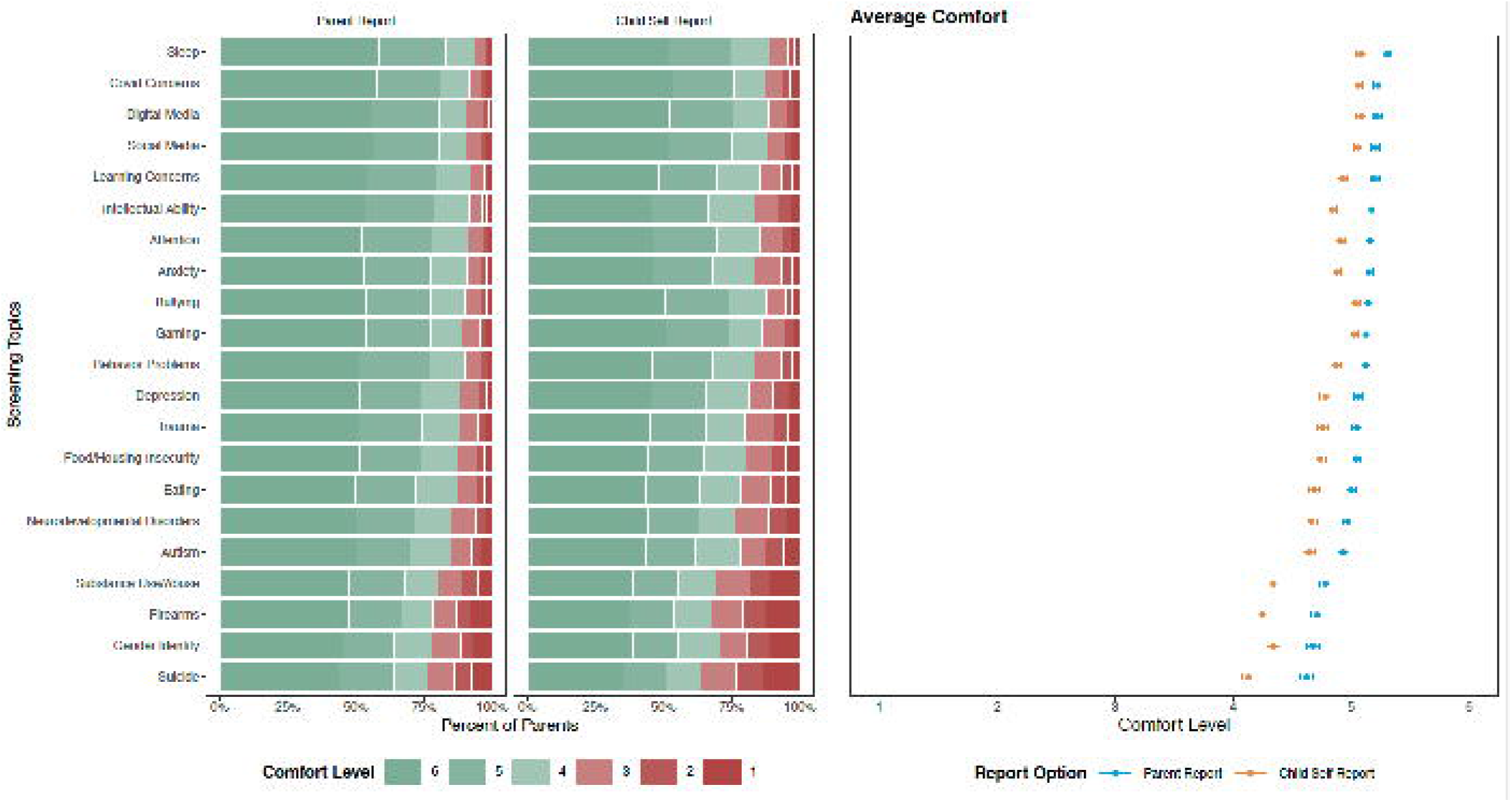
Parental Comfort Levels for Screening Topics by Report Option. Parental comfort levels with parent- and child self-report with specific symptoms and behaviors of 21 topics. Topics are arranged by the average comfort level of parent-reporting in descending order. Comfort levels ranged from 1 to 6, with 6 indicating great comfort. Parents’ comfort levels with parent-reporting were significantly higher than their comfort levels for child self-report for all 21 topics in a series of mixed effect regression models with Benjamini-Hochberg corrections (p<.01-p<.001).

Parental comfort levels for child self-reporting were correlated with their child’s age for all 21 topics, as shown in Figure 4 (*p*<.001 for the topics). Parental comfort levels for 11 of the 21 parent-reporting topics were correlated with the child’s age (substance use/abuse, suicidal ideation, firearms, gender identity, depression, autism, covid-19 concerns, gaming, social media use, digital media use, and food/housing insecurities) (*p*<.05-*p*<.001). When controlling for the parent’s age, these findings remained relatively consistent, with sleep problems, depression and anxiety now also being significantly predicted by the child’s age.

**Figure 4.**
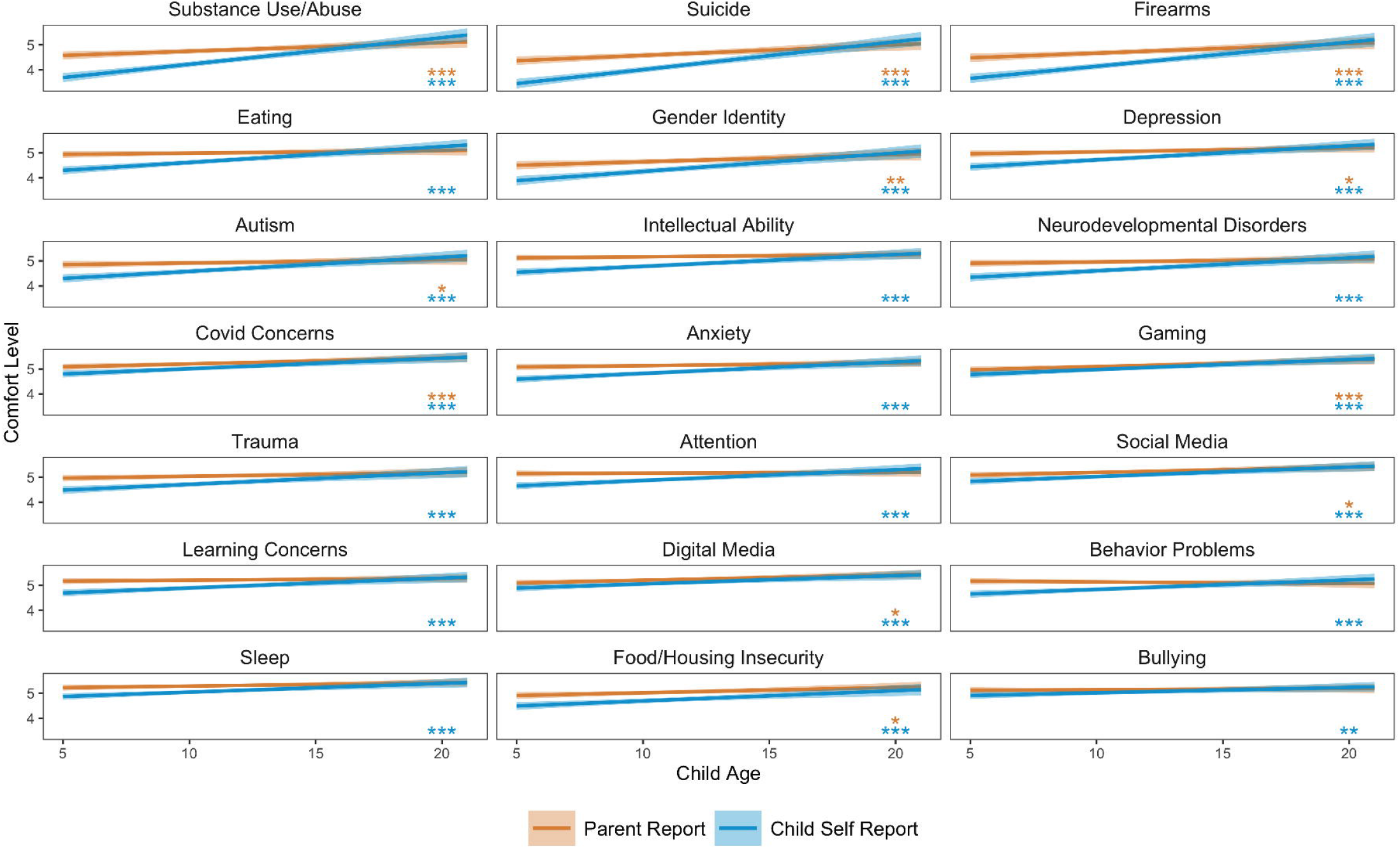
Correlation of Child’s Age and Parents’ Comfort of Screening Topics by Report Option. *\*p<*.*05, **p<*.*01, ***p<*.*001* Results from a series of correlations of child age on parental comfort levels with each report option (parent- and child self-report) for all 21 topics. Topics are ordered by correlation coefficient of child self-report. Statistical significance is denoted by asterisks.

Over 90% of parents agreed (_≥_4 on 6-point Likert scale) that “early detection of problems,” “early intervention,” and “to learn more about my child” were benefits from mental health screening. Other reported benefits included: better “access [to] mental health resources,” “awareness of signs to watch for,” ability “to accommodate/support my child,” “management of symptoms,” and “prevention of problems.”

## DISCUSSION

The present study found that parents were generally comfortable having their child screened for mental health problems, though several preferences were observed. First, parents expressed a clear preference for carrying out screenings on an annual basis - a model that fits well with that of general medical screenings in the primary care setting. Second, parents favored completing the screener in healthcare offices rather than at home, though comfort levels for at-home screening were still relatively high. Third, there was a strong preference for having the interpretation of the findings being provided by physicians and psychologists, with notably lower comfort levels for reviewing results with social workers, office staff or teachers. Regarding screening content, we found that parents’ comfort is highly dependent on screening content and report option (parent-report versus child self-report). Parents were generally comfortable with all 21 screening topics assessed in the present study, though four topics – substance use/abuse, firearms, gender identity, and suicidal ideation – had significantly lower comfort levels. Parents preferred to complete the screeners themselves though they were still relatively comfortable with allowing their child to complete a self-report screener, with their comfort increasing with the child’s age. Finally it is worth noting that our findings were not dependent on country, though some variation in overall comfort levels across countries was present.

Beyond supporting the acceptability of pediatric mental health screening in primary care settings to parents, the present work also suggests potential areas for optimization in future efforts. First, our findings suggest that home-based screenings can minimize workflow interruptions and time costs associated with screening^24,25,29,34,48^ and is an acceptable solution for most parents. As web-based screeners become more widely available, and are integrated into electronic health record systems, healthcare offices may consider this route of administration. This may also allow for increased frequencies of screenings, as certain mental illnesses are known to fluctuate by season,^49^ suggesting that annual screening may not be sufficient for all disorders. Future work should explore the best times of the year to screen youths and whether home-based screening can better capture some of these fluctuations..

Second, parents appear less comfortable with direct screening of their children, than via their own report. Although less concerning for the detection of externalizing disorders, such as ADHD, this can be problematic for the detection of internalizing disorders, such as anxiety and depression. Similarly, some parents expressed decreased comfort with assessment of key topic areas related to risk of harm (e.g., suicidal ideation, substance use, and firearms). Increased efforts towards the education of parents about the potential benefits and risks of screening may help to increase comfort levels for more comprehensive screening processes.

Although focused on primary care settings, the present work included teachers as a reference for assessing preferences for the reporting of results. Our finding of a strong preference for medical professionals may have implications for efforts focused on school-based screening. In particular, they suggest that school-based efforts may benefit from either having a medical or psychological professional on site to have these conversations periodically, or transferring the screening results to the child’s primary care provider for discussion with families.

Study limitations include a requirement that parents be fluent in English and have access to Prolific Academic, an online website. Inclusion of data from multiple countries does suggest some level of generalizability of findings, though it does not exclude potential bias. Current research has not yet addressed cultural and geographic differences in openness to screen, interpret, and take action on pediatric mental health problems and behaviors. Previous studies have suggested that socioeconomic and demographic factors (e.g., race and annual household income) may affect results.^50–52^ Interestingly, the present study did not find a significant influence of these factors on parental comfort levels; however, given the lack of racial diversity in the present sample, this requires further study. Another potential limitation is that the Coronavirus (COVID-19, SARS-CoV-2) pandemic that was ongoing during this survey may have influenced participation rates and responses.

In summary, the present survey demonstrates that there is widespread, cross-national parent acceptability for mental health screening of their offspring, with strong preferences for follow up with experts who can facilitate further evaluation or treatment. This study also illustrates the need to engage both the public who may utilize the benefits of screening, as well as professionals, and some of the key factors (e.g., screening topic, child age, country of residence, and report option) that may enhance the development of future programs to detect and intervene in mental disorders in youth.

## Data Availability

Data will be made available upon request to the authors

## ARTICLE INFORMATION

### Author statement

#### Concept and design

Alexander, Lindsay; Milham, Michael

#### Acquisition, analysis, or interpretation of data

James, Najé; Kass, Mirelle; Moskowitz, Kathleen

#### Drafting of the manuscript

Kass, Mirelle

#### Critical revision of the manuscript for important intellectual content

Alexander, Lindsay; Leventhal, Bennett; Milham, Michael; Merikangas, Kathleen; Salum, Giovanni

#### Statistical analysis

James, Najé; Kass, Mirelle; Moskowitz, Kathleen

#### Administrative, technical, or material support

Alexander, Lindsay

### Supervision

Milham, Michael

## Acknowledgements

The work presented here was primarily supported by a grant award from the Hearst Foundation, as well as gifts to the Child Mind Institute from Phyllis Green, Randolph Cowen, and Joseph Healey, as well as NIMH awards to Dr. Milham (R01MH124045, R01MH091864). This research was also supported by the Intramural Research Program of the NIMH (Merikangas; grant number ZIAMH002953). The funders for this project were not involved in any part of the experimental design, analysis and interpretation of data, or manuscript preparation and submission. Dr. Milham is the Phyllis Green and Randolph Cowen Scholar; Joe Healey provides philanthropic gifts to the Center for the Developing Brain. The views and opinions expressed in this article are those of the authors and should not be construed to represent the views of any of the sponsoring organizations, agencies, or U.S. Government.

## SUPPLEMENTAL TABLES AND FIGURES

**eFigure 1. Parents’ Willingness to Discuss Mental Health and Learning Disorders**.

Parents’ comfort levels on various statements about their willingness and/or ability to discuss mental health and learning disorders. The rating scale was a 6-point Likert scale, with 1=Disagree and 6=Agree.

**eFigure 2. Average Parental Comfort Levels by Country**.

* *p*<.05, \*\**p*<.01, \*\*\**p*<.001.

Parental comfort levels of parent report and child self-report for various screening topics by country sample (US: *n*=265; UK: *n*=282; Canada: *n*=171; Other Countries: *n*=254). Topics are ordered according to the entire sample (*N*=972). Comfort levels ranged from 1 (not comfortable) to 6 (comfortable).

