## Supplementary figures and images for "Robust Parental Preferences in Mental Health Screening in Youth From a Multinational Online Survey"

### Supplemental Figure 1

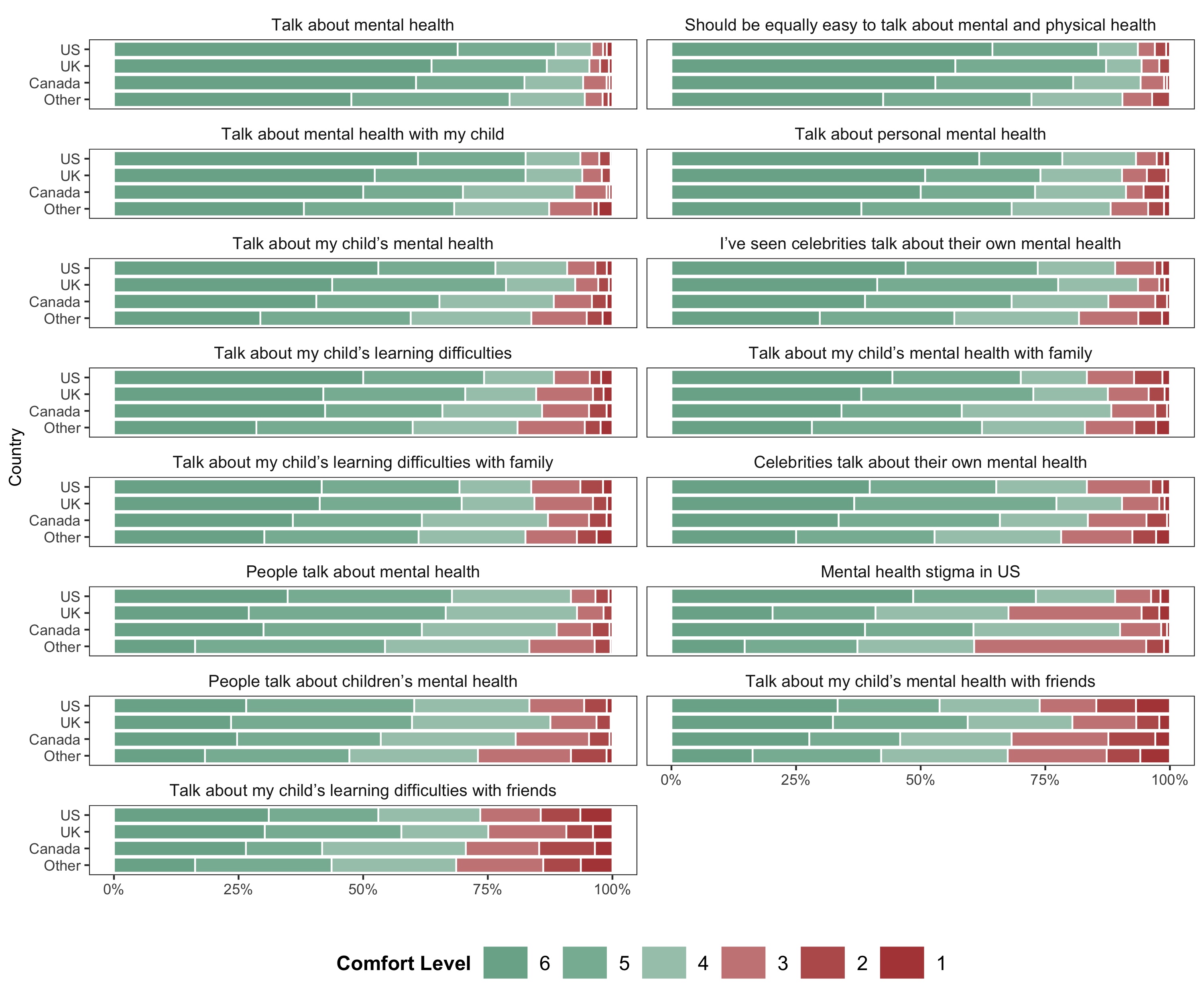

### Supplemental Figure 2

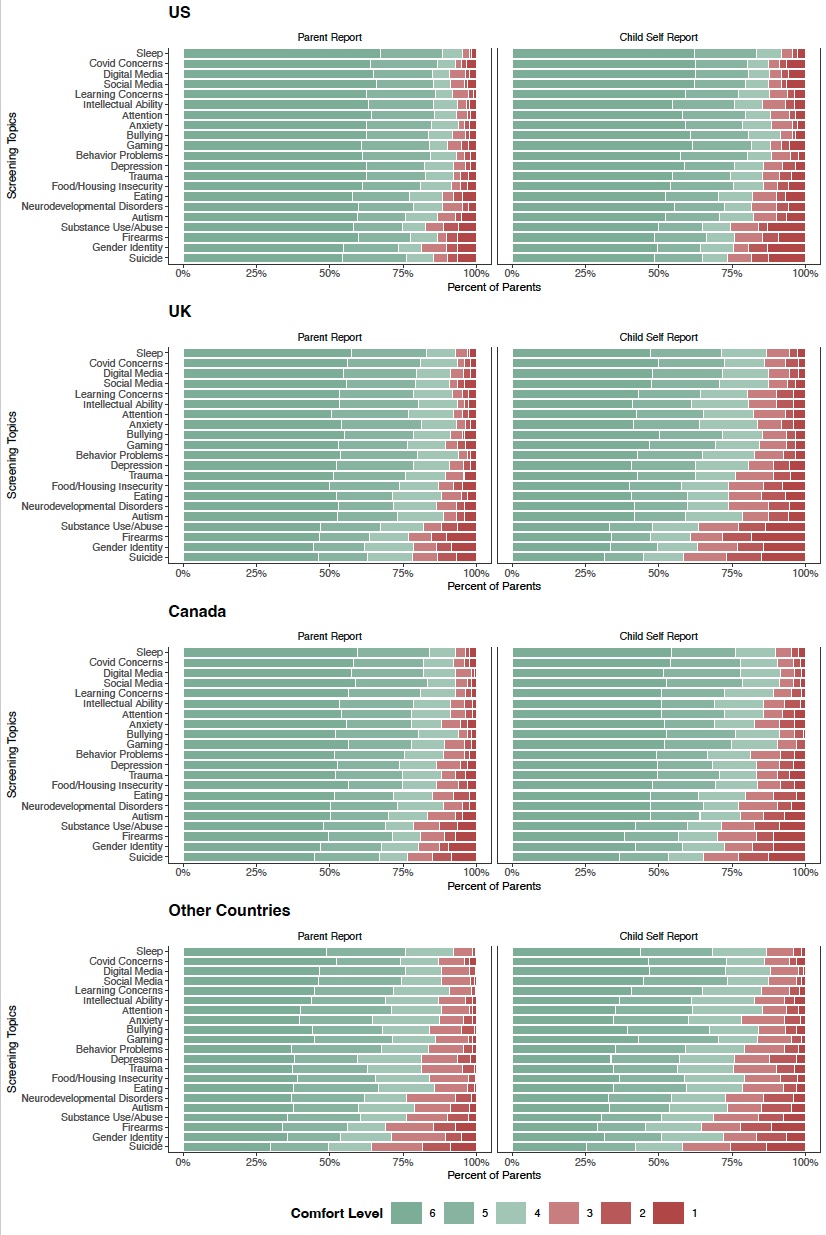
